## Supplementary data 1-5 for "Epidemiological and immunological features of obesity and SARS-CoV-2"

**Nilles EJ, et al.**

### Supplemental data 1: Immunoassays

#### ELISA IgG assay

Serological analyses were performed using Ragon/MGH enzyme-linked immunosorbent assay that detects IgG against the receptor binding domain (RBD) of the SARS-CoV-2 spike glycoprotein (provided by Aaron Schmidt) using a previously described method [1]. Briefly, 384-well plates were coated with 0·5ug/ml of RBD for 1h at 37$^{o}$C. The plates were then blocked with BSA containing buffer, washed, and plasma samples added at a 1:100 dilution in duplicate for 1h at 37$^{o}$C, washed and then detected with a secondary anti-IgG (Bethyl Laboratories). The secondary was washed away after 1h, and the colormetric detector was added (Thermo Fisher) for 5 mins, the reaction was stopped, and the luminescence was acquired on a luminometer at an absorbance of 450/540nm. A positive cutoff was equal to the mean of the OD-converted µg/ml values of the negative control wells on the respective plate plus five times the standard deviation of the concentration from over 100 pre-COVID-19 plasma samples. The background-corrected concentrations were divided by the cutoff to generate signal-to-cutoff (S/CO) ratios. Assay performance has been externally validated in a blinded fashion at 99·6% specific and benchmarked against commercial EUA approved assays.[2]

#### IgG subclass, isotype and FcγR binding

SARS-CoV2 specific antibody subclass and isotypes, and FcγR binding was analyzed using a custom Luminex multiplexed assay. SARS2-CoV2-RBD, SARS2-CoV2-N and SARS2-CoV2-S were coupled to magnetic Luminex beads (Luminex Corp, TX, USA) by carbodiimide-NHS ester-coupling (Thermo Fisher). Dilution curves were performed on pooled samples from the cohort to determine dilutions in the linear range for each detection reagent. Coupled beads were then incubated with different plasma dilutions (between 1:100 and 1:1,000 depending on the secondary reagent) for 2 hours at room temperature in 384 well plates (Greiner Bio-One, Germany). Unbound antibodies were washed away and IgG1, IgG3, IgM or IgA1 were detected with their respective PE-conjugated antibody (all polyclonal, Southern Biotech, AL, USA). For the FcγR3b binding, a PE-Streptavidin (Agilent Technologies, CA, USA) coupled recombinant biotinylated human FcγR3b protein (Duke Protein Production Facility) was used as a secondary probe. After 1 h incubation, excessive secondary reagent was washed away and the relative antibody concentration per antigen determined on an IQue analyzer (IntelliCyt, NM, USA). Samples with signals 5-times the standard deviation of the PBS-control well were considered as positive.

**SARS-CoV2 antibody mediated virus neutralization**

The ability of antibodies to neutralize virus was assessed on a 2019-nCoV pseudovirus neutralization assay, as described previously [3]. In brief, HEK293T cells were transfected with pcDNA3·1(-)-hACE2 (Addgene). 12 hours post transfection; the HEK293T/hACE2 cells were seeded in 96-well plates (2 104 cells/well) and incubated overnight. Heat (56°C, 30 min) inactivated plasma samples were serially diluted and mixed with 50µl of pseudoviruses, incubated at 37°C for 1 hour and added to the HEK293T/hACE2 cells. Forty-eight hours after infection, cells were lysed in Steady-Glo Luciferase Assay detection (Promega). A standard quantity of cell lysate was used in the luciferase assay with luciferase assay reagent (Promega) according to the manufacturer's protocol. SARS-CoV-2 neutralization titers were defined as the sample dilution at which a 50% reduction in RLU was observed relative to the average of the virus control wells.

**ELISPOT**

PBMCs were isolated and frozen from EDTA blood within 24 hours after collection using Sepmate tubes (Stemcell Technology). PVDV membrane plates (Millipore, MA, USA) were coated with anti-human IFNγ antibody (clone: 1-DK1, conc.: 2 µg/ml) overnight. Previously frozen and overnight rested PBMC samples were counted and 2 X $10^5$ PBMCs were added per well with S or N overlapping peptide pools (both Miltenyi, Germany) at 1·25 µg/ml peptide, overnight. Medium alone was used as a negative control. Pools of 23 MHC-I restricted peptides from human Cytomegalovirus, Eppstein Barr virus and Influenza virus (CEF, Anaspec Inc.) and 35 MHC-II restricted peptides from human Cytomegalovirus, Epstein Barr virus, Influenza virus, Tetanus toxin and Adenovirus 5 (CEFTA, Mabtech Inc.) were used as positive controls. IFNγ secretion was detected with a biotinylated anti-human IFNγ antibody (clone: 7 B6-1) and ALP conjugated-Streptavidin. Spots were developed with 1-Step BCIP/NBT-plus reagent (Mabtech Inc.) for 20 minutes. Membranes were dried and spots were analyzed and counted on an ImmunoSpot CTL analyzer. A response was considered positive only if there were $\geq$ 25 SFCs/$10^6$ PBMC.

**Supplementary data 2. Characteristics and serostatus of South Texas site with high seropositive rate (22·5%) (N=712)**

| Characteristic1 | All participants (n=712) | Seropositive participants (n=160) | | OR (95% CI) | P-Value2 |
|---|---|---|---|---|---|
| | N | N | % | | |
| **Age group** | | | | | |
| **18-29 y** | 345 | 86 | 24.9% | ref | |
| **30-39 y** | 181 | 35 | 19.3% | 0.72 (0.46 to 1.12) | 0.1488 |
| **40-49 y** | 128 | 28 | 21.9% | 0.84 (0.52 to 1.37) | 0.4907 |
| **50-59 y** | 45 | 10 | 22.2% | 0.86 (0.41 to 1.81) | 0.6921 |
| **60+ y** | 12 | 1 | 8.3% | 0.27 (0.03 to 2.15) | 0.2179 |
| **Body mass index** | | | | | |
| **<18.5** | 8 | 1 | 12.5% | 0.35 (0.04 to 2.91) | 0.3291 |
| **18.5-<25** | 144 | 42 | 29.2% | ref | |
| **25-<30** | 230 | 53 | 23.0% | 0.73 (0.45 to 1.17) | 0.1864 |
| **30-<35** | 162 | 34 | 21.0% | 0.65 (0.38 to 1.09) | 0.0995 |
| **35-<40** | 63 | 14 | 22.2% | 0.69 (0.35 to 1.39) | 0.3022 |
| **≥40** | 22 | 1 | 4.5% | 0.12 (0.02 to 0.89) | 0.0380* |
| **Ethnicity** | | | | | |
| **Not Hispanic/Not Latinx** | 93 | 16 | 17.2% | ref | |
| **Hispanic/Latinx** | 493 | 117 | 23.7% | 1.50 (0.84 to 2.67) | 0.1702 |
| **Race** | | | | | |
| **White** | 375 | 82 | 21.9% | ref | |
| **American Indian/Alaska Native** | 9 | 3 | 33.3% | 1.79 (0.44 to 7.30) | 0.4190 |
| **Asian** | 9 | 3 | 33.3% | 1.79 (0.44 to 7.30) | 0.4190 |
| **Black** | 7 | 1 | 14.3% | 0.60 (0.07 to 5.02) | 0.6336 |
| **Native Hawaiian/Pacific Islander** | 3 | 0 | 0.0% | 0.00 (0.00 to Inf) | 0.9792 |
| **More than one race** | 26 | 4 | 15.4% | 0.65 (0.22 to 1.94) | 0.4393 |
| **Sex** | | | | | |
| **Female** | 44 | 10 | 22.7% | ref | |
| **Male** | 634 | 144 | 22.7% | 1.00 (0.48 to 2.07) | 0.9982 |
| **Children ≤ 18 y in household** | | | | | |
| **No** | 407 | 102 | 25.1% | ref | |
| **Yes** | 273 | 54 | 19.8% | 0.74 (0.51 to 1.07) | 0.1091 |
| **No. in household** | | | | | |
| **1** | 98 | 22 | 22.4% | ref | |
| **2-4** | 393 | 97 | 24.7% | 1.13 (0.67 to 1.92) | 0.6446 |
| **>4** | 173 | 34 | 19.7% | 0.84 (0.46 to 1.55) | 0.5852 |
| **Comorbidity3** | | | | | |
| **Asthma** | 27 | 4 | 14.8% | 0.59 (0.20 to 1.73) | 0.3364 |
| **Hypertension** | 47 | 11 | 23.4% | 1.06 (0.53 to 2.13) | 0.8741 |
| **Diabetes mellitus** | 15 | 3 | 20.0% | 0.86 (0.24 to 3.09) | 0.8168 |
| **Coronary heart disease (I got zero** | 3 | 1 | 33.3% | | |
| **Other lung disease** | 2 | 1 | 50.0% | 3.47 (0.22 to 55.71) | 0.3805 |
| **Other chronic medical** | 9 | 2 | 22.2% | 0.99 (0.20 to 4.79) | 0.9856 |
| **Smoking history** | | | | | |
| **Never** | 546 | 124 | 22.7% | ref | |
| **Prior** | 58 | 15 | 25.9% | 1.18 (0.64 to 2.20) | 0.5933 |
| **Current** | 80 | 17 | 21.3% | 0.92 (0.52 to 1.62) | 0.7641 |

1 Not reported data: age group (n=1), BMI (83), ethnicity (126), race (283), sex (34), children in HH (31), No. in HH (47), comorbidities (28).

2 * P<0.05

3 Other comorbidities with no seropositive individuals (n= number reporting comorbidity): chronic kidney disease (2), stroke (2), COPD (0), Heart failure (0), cancer receiving treatment (1), cancer not receiving treatment (3), other heart disease (0), other immunocompromised (0).

**Supplementary data 3. Symptoms reported by healthy/normal versus overweight but not obese seropositive individuals (n=178)**

| Clinical feature | Normal weight (18.5-<25) (n=89) | | Overweight (25-<30) (n=89) | | P-value1 |
|---|---|---|---|---|---|
| | N | % | N | % | |
| Fever | 7 | 7.9% | 6 | 6.7% | 0.76 |
| Chills | 9 | 10.1% | 11 | 12.4% | 0.63 |
| Cough | 16 | 18.0% | 14 | 15.7% | 0.68 |
| Loss of smell | 9 | 10.1% | 14 | 15.7% | 0.27 |
| Loss of taste | 8 | 9.0% | 11 | 12.4% | 0.46 |
| Nausea and/or vomiting | 5 | 5.6% | 5 | 5.6% | 1.00 |
| Diarrhea | 7 | 7.9% | 11 | 12.4% | 0.32 |
| Congestion | 18 | 20.2% | 17 | 19.1% | 0.85 |
| Sore throat | 11 | 12.4% | 14 | 15.7% | 0.54 |
| Myalgias | 13 | 14.6% | 11 | 12.4% | 0.67 |
| Increased fatigue | 23 | 25.8% | 14 | 15.7% | 0.10 |
| **Aggregate symptom measures2** | | | | | |
| Any symptom | 36 | 40.4% | 35 | 39.3% | 0.88 |
| ≥3 symptoms | 19 | 21.3% | 20 | 22.5% | 0.85 |
| ≥6 symptoms | 11 | 12.4% | 8 | 9.0% | 0.46 |
| Any primary symptom | 20 | 22.5% | 22 | 24.7% | 0.73 |
| ≥3 primary symptoms | 10 | 11.2% | 10 | 11.2% | 1.00 |
| Symptoms count (mean) | 126 | 1.42 | 128 | 1.44 | 0.96 |
| Primary symptom count (mean) | 49 | 0.55 | 56 | 0.63 | 0.68 |

1 Chi-squared test of proportions and ANOVA of means

2 Primary symptoms: Fever, chills, cough, loss of smell and loss of taste

**Supplementary data 4. Symptom reporting by age group and obesity status among SARS-CoV-2 seropositive individuals.** (A) Heatmap shows consistently higher symptom reporting amongst obese individuals in the 19-29- and 29-39-year age groups but not ≥40-year age group. Number of individuals in each category are listed below obesity markers. (B) Table lists relevant values. * indicates P<0·05 for difference between obese and non-obese in that age category with Chi-squared test for proportions and ANOVA for test of mean.

A

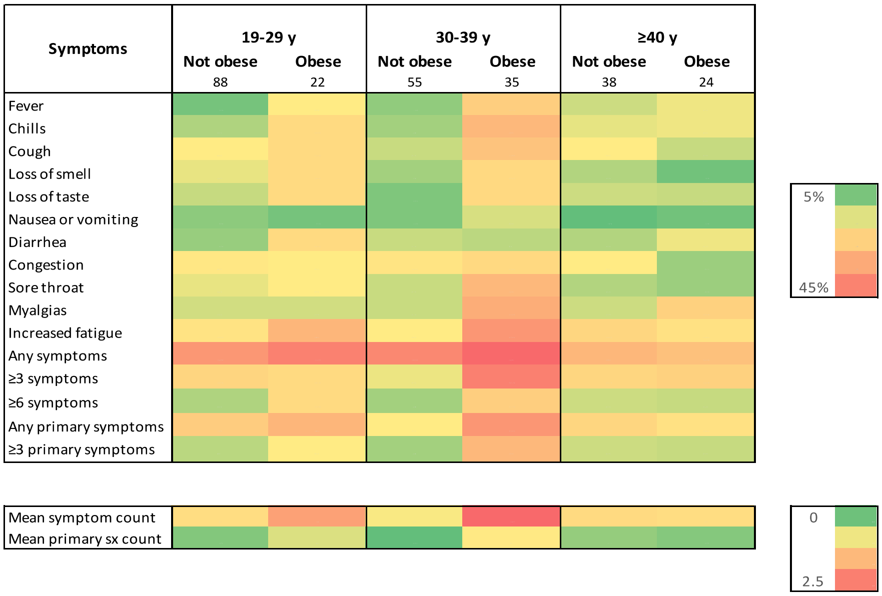


B

| Symptoms | 19-29 y Not obese (88) | 19-29 y Obese (22) | 30-39 y Not obese (55) | 30-39 y Obese (35) | ≥40 y Not obese (38) | ≥40 y Obese (24) |
|---|---|---|---|---|---|---|
| Fever | 4.5% | 18.2%* | 7.3% | 25.7%* | 13.2% | 16.7% |
| Chills | 10.2% | 22.7% | 9.1% | 31.4%* | 15.8% | 16.7% |
| Cough | 18.2% | 22.7% | 12.7% | 28.6% | 18.4% | 12.5% |
| Loss of smell | 15.9% | 22.7% | 9.1% | 22.9% | 10.5% | 4.2% |
| Loss of taste | 12.5% | 22.7% | 5.5% | 22.9%* | 13.2% | 12.5% |
| Nausea or vomiting | 6.8% | 4.5% | 5.5% | 14.3% | 2.6% | 4.2% |
| Diarrhea | 8.0% | 22.7%* | 12.7% | 11.4% | 10.5% | 16.7% |
| Congestion | 19.3% | 18.2% | 20.0% | 22.9% | 18.4% | 8.3% |
| Sore throat | 15.9% | 18.2% | 12.7% | 31.4%* | 10.5% | 8.3% |
| Myalgias | 13.6% | 13.6% | 12.7% | 34.3%* | 13.2% | 25.0% |
| Increased fatigue | 20.5% | 31.8% | 18.2% | 40.0%* | 23.7% | 20.8% |
| Any symptoms | 39.8% | 45.5% | 43.6% | 51.4% | 31.6% | 29.2% |
| ≥3 symptoms | 23.9% | 22.7% | 16.4% | 45.7%* | 23.7% | 25.0% |
| ≥6 symptoms | 10.2% | 22.7% | 9.1% | 25.7%* | 13.2% | 12.5% |
| Any primary symptoms | 26.1% | 31.8% | 18.2% | 40.0%* | 23.7% | 20.8% |
| ≥3 primary symptoms | 11.4% | 18.2% | 9.1% | 31.4%* | 13.2% | 12.5% |
| Mean symptom count | 1.45 | 2.18 | 1.25 | 2.86* | 1.50 | 1.46 |
| Mean primary sx count | 0.61 | 1.09 | 0.44 | 1.31* | 0.71 | 0.63 |

**Supplementary data 5.** Results of univariate differences (Mann-Whitney U test) in immune feature by obese (n=25) versus non-obese (n=52) status

| **Feature** | **p-value** |
|---|---|
| FcgR3b-RBD | 2·84E-01 |
| FcgR3b-S | 2·30E-01 |
| FcgR3b-N | 1·70E-01 |
| FcgR3b-NTD | 2·47E-01 |
| IgA1-N | 3·14E-01 |
| IgA1-S | 1·41E-01 |
| IgA1-NTD | 3·32E-01 |
| IgA1-RBD | 2·38E-01 |
| IgG1-N | 1·33E-01 |
| IgG1-NTD | 1·09E-01 |
| IgG1-S | 2·23E-01 |
| IgG1-RBD | 3·20E-01 |
| IgG3-N | 3·51E-01 |
| IgG3-S | 4·38E-01 |
| IgG3-NTD | 4·24E-01 |
| IgG3-RBD | 1·51E-01 |
| IgM-RBD | 2·23E-01 |
| IgM-S | 2·03E-01 |
| IgM-NTD | 2·84E-01 |
| IgM-N | 1·45E-01 |
